# Serum Neutralizing Activity of mRNA-1273 Against the SARS-CoV-2 B.1.1.529 (Omicron) Variant: A Preliminary Report

**DOI:** 10.1101/2022.01.28.21268247

**Authors:** Diana Lee, Laura E. Avena, Daniela Montes Berrueta, Matthew Koch, Angela Choi, Judy Oestreicher, William Hillebrand, Honghong Zhou, Rolando Pajon, Andrea Carfi, Darin Edwards, Kai Wu

## Abstract

The emergence of the severe acute respiratory syndrome coronavirus 2 (SARS-CoV-2) B.1.1.529 (Omicron) variant has led to growing concerns of increased transmissibility and escape of both natural and vaccine-induced immunity. In this analysis, sera from adult participants in a phase 2 clinical study (NCT04405076) were tested for neutralizing activity against B.1.1.529 after a 2-dose (100 µg) mRNA-1273 primary vaccination series and after a 50-µg mRNA-1273 booster dose. Results from this preliminary analysis show that 1 month after completing the primary series, mRNA-1273-elicited serum neutralization of B.1.1.529 was below the lower limit of quantification; however, neutralization was observed at 2 weeks after the mRNA-1273 booster dose, although at a reduced level relative to wild-type SARS-CoV-2 (D614G) and lower than that observed against D614G at 1 month after the primary series.

## INTRODUCTION

Throughout the coronavirus disease 2019 (COVID-19) pandemic, severe acute respiratory syndrome coronavirus 2 (SARS-CoV-2) has continuously evolved into several variants of interest (VOIs) and variants of concern (VOCs) [1], altering its pathogenic potential and ability to evade the immune system. Most recently, the emergence of the B.1.1.529 (Omicron) variant has raised substantial public health concerns, with the World Health Organization (WHO) designating B.1.1.529 as a VOC on November 26, 2021 [2]. With multiple mutations in the spike (S) glycoprotein, including but not limited to several identified in other SARS-CoV-2 VOCs that were linked to increased transmissibility and reduced susceptibility to antibody neutralization [3], B.1.1.529 is under intense international scrutiny for its potential to evade both natural and vaccine-induced immunity [2, 4-8].

The mRNA-1273 vaccine (SPIKEVAX™, Moderna Inc., Cambridge, MA, USA), a lipid nanoparticle-encapsulated mRNA-based vaccine encoding the S glycoprotein of the SARS-CoV-2 Wuhan-Hu-1 isolate, has been shown effective against symptomatic and severe COVID-19 in clinical and real-world settings [9-11], including against B.1.617.2 (Delta) and B.1.351 (Beta) variants [12, 13]. Further, mRNA-1273 induces neutralizing antibodies against wild-type SARS-CoV-2 and several previously identified VOCs and VOIs, although certain variants, notably B.1.351 (Beta), B.1.617.2 (Delta), and P.1 (Gamma), showed reduced neutralizing antibody titers compared to wild-type (D614G) virus [14, 15].

As serum neutralizing antibody levels are highly predictive of immune protection from symptomatic SARS-CoV-2 infection [16, 17], we assessed the neutralization activity of mRNA-1273 sera against B.1.1.529 after completion of the 2-dose (100 µg) primary vaccination series and after a 50-µg booster dose.

## MATERIALS AND METHODS

### Participant samples

Sera were collected from healthy adult participants enrolled in the open-label interventional phase of a phase 2 clinical study of mRNA-1273 (NCT04405076) [18]. Sera from matched participants (n = 10) were collected 1 month (Day 57) after completing the 2-dose (100 µg) primary series and 2 weeks after a single 50-µg booster dose. Sera collected from an additional 10 participants 2 weeks after a single 50-µg booster dose were also evaluated. All participants were randomly selected for inclusion in this preliminary analysis based on visit assessments completed and sample availability of pre-booster sera. All participants provided informed consent and all study materials were approved by a central institutional review board (Advarra; Columbia, Maryland); the study was conducted in accordance with the International Council on Harmonization of Good Clinical Practice guidelines.

### Recombinant vesicular stomatitis virus (VSV)-based pseudovirus assay

The codon-optimized full-length S protein of the original Wuhan-Hu-1 SARS-CoV-2 isolate with D614G mutation was cloned into a pCAGGS vector [14]. The SARS-CoV-2 B.1.1.529 variant was then generated using site-directed mutagenesis of the codon-optimized D614G vector to incorporate the S mutations listed in **Table 1**. As previously described [19], the SARS-CoV-2 full-length S-pseudotyped recombinant vesicular stomatitis virus ΔG (VSVΔG)–firefly luciferase virus was made by first transfecting BHK-21/WI-2 cells (Kerafast) with the S-expression plasmid and then subsequently infecting cells with the VSVΔG-firefly luciferase virus. The research grade neutralization assay followed a previously described protocol [14]. In brief, pseudovirus was first mixed with serially diluted serum samples and incubated at 37 °C for 45 minutes. This pseudovirus–serum mix was then used to infect A549-hACE2-TMPRSS2 cells [20] for 18 hours at 37 °C, followed by the addition of ONE-Glo reagent (Promega) to measure luciferase signal by relative luminescence units (RLUs). Percentage of neutralization was calculated based on the RLUs of the virus-only control, then analyzed using 4-parameter logistic curve in Prism v.9 (GraphPad Software, Inc.).

**Table 1.** Mutations in the spike protein of the SARS-CoV-2 B.1.1.529 (Omicron) variant evaluated in this study.

| Variant name | WHO nomenclature | Region identified | Mutations in spike protein |
| --- | --- | --- | --- |
| **D614G** | - | Wild-type<br>global variant | D614G |
| **B.1.1.529** | Omicron | South Africa | A67V, Δ69-70, T95I, G142D/Δ143-145, Δ211, L212I, ins214EPE, G339D, S371L, S373P, S375F, K417N, N440K, G446S, S477N, T478K, E484A, Q493R, G496S, Q498R, N501Y, Y505H, T547K, D614G, H655Y, N679K, P681H, N764K, D796Y, N856K, Q954H, N969K, L981F |

WHO, World Health Organization.

### Statistical Analysis

A 2-sided Wilcoxon matched-pairs signed rank test was utilized to compare matched patients against the wild-type (D614G) and B.1.1.529 viruses. Geometric mean titers (GMTs) and lower limit of quantification (LLOQ) were included. Statistical analyses were performed in Prism v.9.

## RESULTS

Neutralization activity of mRNA-1273 sera against B.1.1.529 SARS-CoV-2 variant (Omicron) pseudovirus was assessed in 10 adult participants (9 participants were ≥55 years of age, of which 6 were ≥65 years of age) after completing the 2-dose (100 µg) primary series and in 20 adult participants (19 participants were aged ≥55 years, of which 11 were aged ≥65 years) after receiving a 50-µg booster dose. At 1 month after the primary series, neutralizing antibodies (NAbs) against B.1.1.529 were below the LLOQ and significantly lower than those against the wild-type (D614G) virus (*P*=0.002; **Figure 1**). At 2 weeks after the mRNA-1273 booster dose, NAbs against B.1.1.529 increased with the majority of sera from mRNA-1273 vaccine recipients showing neutralization activity against the variant; however, NAb titers against B.1.1.529 were reduced compared to the wild-type (D614G) virus (*P*<0.0001; **Figure 1**). Notably, neutralization titers against B.1.1.529 after the mRNA-1273 booster dose were also lower than those against the wild-type (D614G) virus after the primary series (**Figure 1**).

**Figure 1.**
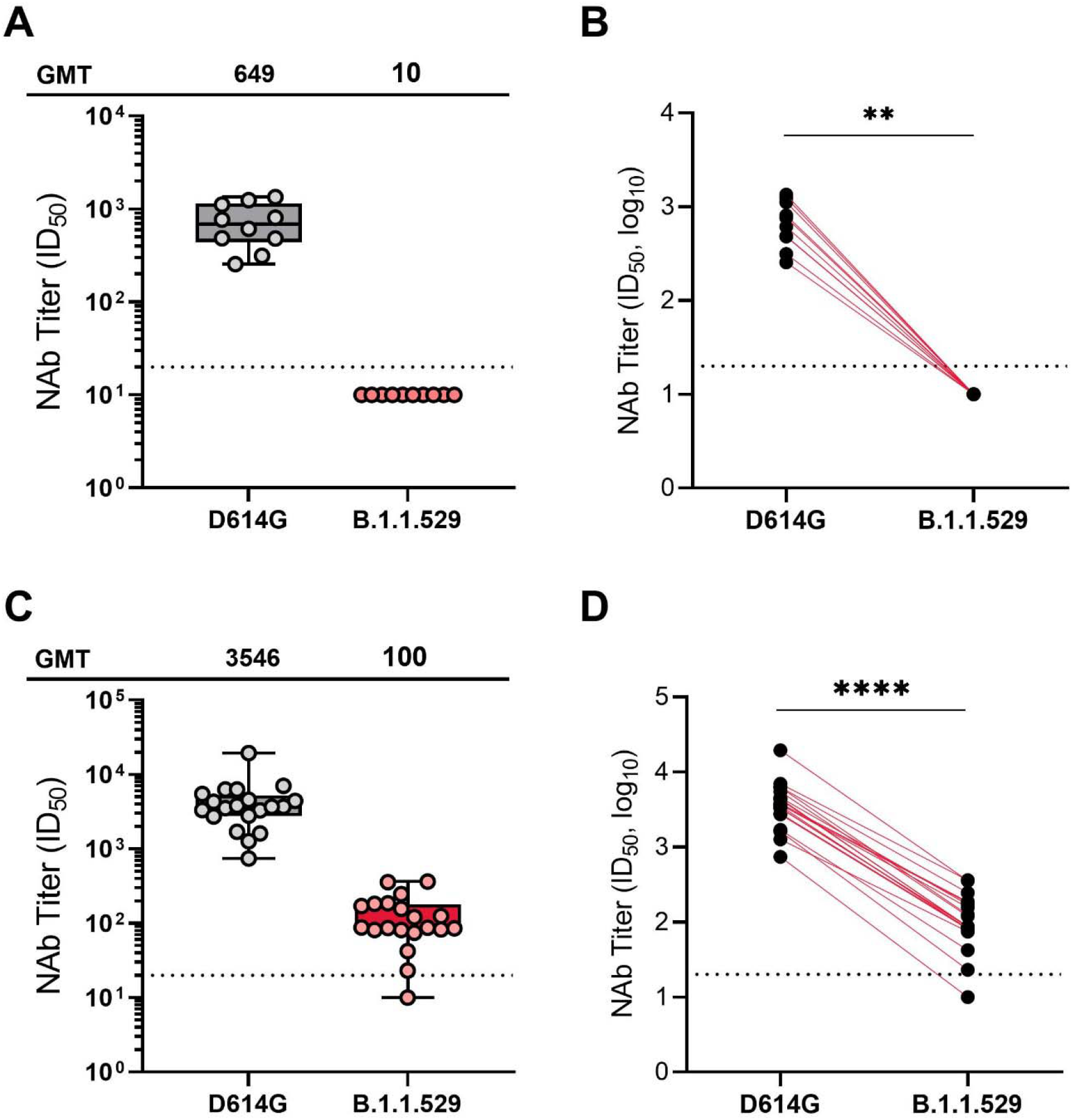
Neutralization of SARS-CoV-2 pseudoviruses in serum samples at 1 month post mRNA-1273 primary series (A and B) and 2 weeks post mRNA-1273 booster (C and D). Serum samples were obtained from participants enrolled in the mRNA-1273 vaccine phase 2 open label study wherein participants who received a 2-dose primary series (100 µg) during the double-blind phase of the trial received a single booster dose of mRNA-1273 (50 µg). For Panels A and B, samples were collected at Day 57 (1 month following the primary series) and for panels C and D, samples were collected at day 15 (2 weeks following the booster dose). A recombinant vesicular stomatitis virus–based pseudovirus neutralization assay was used to measure neutralization. The pseudoviruses tested incorporated D614G (WT) or the spike substitutions present in B.1.1.529 (Omicron). The reciprocal neutralizing titers on the pseudovirus neutralization assay at ID_50_ are shown. In panels A and C, boxes and horizontal bars denote the IQR range and the GMT, respectively. Whisker end points are equal to the values below or above the median at 1.5 times the IQR. In panels B and D, the colored lines connect the D614G and B.1.1.529 neutralization titers in matched samples. A 2-tailed Wilcoxon matched-pairs signed-rank test was performed (**, *P*<0.01; ****, *P*<0.0001). In all panels, the dots represent results from individual serum samples, and the dotted line represents the lower limit of quantification for titers at 20 ID_50_. GMT, geometric mean titer; ID_50_, 50% inhibitory dilution; IQR, interquartile range; NAb, neutralizing antibody; WT, wild-type.

## DISCUSSION

In this preliminary report, mRNA-1273–elicited neutralization of B.1.1.529 was measured in a research-grade VSV-based SARS-CoV-2 pseudovirus neutralization assay in sera collected from phase 2 clinical trial participants 1 month after completing a 2-dose primary series of mRNA-1273 (100 µg; n = 10 participants, of which 9 were aged ≥55 years) or 2 weeks after a booster dose (50 µg; n = 20 participants, of which 19 were aged ≥55 years). Robust neutralization of the wild-type virus (D614G) was measured both after the primary series and after the booster dose. In contrast, mRNA-1273–elicited neutralization against B.1.1.529 was below the assay lower limit of quantification after the 2-dose primary series and was significantly reduced relative to the wild-type (D614G) virus 2 weeks after the booster dose. mRNA-1273–elicited neutralization against B.1.1.529 after the booster dose remained lower than that observed against the wild-type (D614G) virus 1 month after the primary series.

Although a substantial reduction in NAb titers against B.1.1.529 compared to wild-type (D614G) virus was observed in this analysis, it is important to note that NAb titers against B.1.1.529 after the booster dose were still above levels previously predicted to provide protection against severe infection [16]. These predictions are supported by findings based on the VSV assay showing that neutralizing titers against the Delta variant 6 months after primary mRNA-1273 vaccination were below 100 (GMT) [21], yet recently published real-world studies have shown high vaccine effectiveness up to 8 months after 2 doses of mRNA-1273 in a time when Delta predominated [12, 22].

These preliminary findings should be considered with the known limitations of this analysis, including the small sample size that comprised primarily older participants and may not reflect the range of NAb titers observed across different age and demographic groups. Moreover, the assay used in this analysis was research-grade but had limited optimization against the B.1.1.529 virus prior to this analysis [21]. Additional studies assessing durability of NAb titers against B.1.1.529 after an mRNA-1273 booster dose and corroboration of these results with real-world effectiveness data are needed.

These findings are consistent with a growing number of reports indicating that COVID-19 vaccines have reduced neutralizing activity against the B.1.1.529 variant [2, 4-8, 23], thus further emphasizing the need for a booster dose. Additional evaluation of vaccine-induced immunity among a representative population, as well as an understanding of the clinical characteristics of this newly-identified VOC, are required. Further insight should be gained by an upcoming analysis of a broad panel of sera collected from mRNA-1273 vaccine recipients 1 month after the primary series and 1 month after an mRNA-1273 booster dose (50 µg or 100 µg) using both live virus and pseudovirus neutralization assays. These data can allow for direct corroboration of neutralization results and provide more comprehensive information on mRNA-1273 vaccine–induced immunity against the SARS-CoV-2 B.1.1.529 variant.

## Data Availability

The authors declare all data supporting the findings of this analysis are available within this article.

## ACKNOWLEDGMENTS

This work used samples from a phase 2 mRNA-1273 study (NCT04405076) that was supported in part with federal funds from the Office of the Assistant Secretary for Preparedness and Response, Biomedical Advanced Research and Development Authority, under Contract No. 75A50120C00034, and Moderna, Inc. We thank Dr. Elisa Russo and Dr. Michael Whitt for kind support on recombinant VSV-based SARS-CoV-2 pseudovirus production. Medical writing and editorial assistance were provided by Emily Stackpole, PhD and Srividya Ramachandran, PhD, of MEDiSTRAVA in accordance with Good Publication Practice (GPP3) guidelines, funded by Moderna Inc., and under the direction of the authors.

